# Minimal Residual Disease via circulating tumor DNA Predicts Exceptional Response in HER2-Positive Metastatic Breast Cancer

**DOI:** 10.64898/2026.07.09.26357137

**Authors:** Stefania Morganti, Catherine Song, Ningxuan Zhou, Katherine Santos, Pia Jain, Laurel Walsh, Rachel Li, Justin Rhoades, Katie Gilligan, Gregory Kirkner, Catherine Stever, Ashka Patel, Melissa E. Hughes, Nolan Priedigkeit, G. Mike Makrigiorgios, Ian E. Krop, Giuseppe Curigliano, Eric P. Winer, Sara M. Tolaney, Nabihah Tayob, Hillary Heiling, Kan Xiong, Nancy U. Lin, Viktor A. Adalsteinsson, Heather A. Parsons

**Affiliations:** Breast Oncology Program, Dana-Farber Cancer Institute, Boston, MA, USA; Harvard Medical School, Boston, MA, USA; Gerstner Center for Cancer Diagnostics, Broad Institute of MIT and Harvard, Cambridge, MA, USA; Department of Data Science, Dana-Farber Cancer Institute, Boston, MA, USA; Cancer Program, Broad Institute of MIT and Harvard, Cambridge, MA, USA; Department of Radiation Oncology, Dana-Farber Cancer Institute, Boston, MA, USA; Yale Cancer Center, New Haven, CT, USA; European Institute of Oncology, IRCCS, Milano, Italy; Department of Oncology and Hemato-Oncology, University of Milano, Milano, Italy

## Abstract

**Purpose:** Exceptional responses are frequent in patients with HER2-positive (HER2+) metastatic breast cancer (MBC), but predictive biomarkers are lacking. We aimed to investigate the association between detection of minimal residual disease (MRD) via circulating tumor DNA (ctDNA) and exceptional response to first-line HER2 targeted therapy for MBC.

**Patients and Methods:** We identified exceptional (real-world progression-free survival [rwPFS] ≥3 years) and conventional (rwPFS <3 years) responders treated with first-line HER2 targeted therapy for HER2+ MBC and plasma collected at landmark timepoints (e.g., baseline, year [Y] 1, Y2, Y3, at progression). We generated personalized, tissue-informed MRD assays using MAESTRO mutation enrichment sequencing in a pooled format. The primary endpoint was the association between MRD status at Y1 and rwPFS.

**Results:** Of 70 patients, 63 (90%) (40 exceptional and 23 conventional responders) had sufficient samples and successful assay design; MAESTRO was run on 149 samples. A median of 1,823 (range 387-5,000) tumor-specific mutations were tracked per patient. MRD was detected in 49 (32%) samples (median tumor fraction [TFx] 936 ppm; range 3.8-164,068 ppm); 15 (31%) samples had TFx <100 ppm. MRD was associated with outcomes: 0/27 [0%] exceptional versus 9/12 [75%] conventional responders (p<0.001) had detectable MRD at Y1. Exceptional responders who remained progression-free were always MRD-negative (n=30) or cleared MRD by Y1 (n=3). Six exceptional responders experienced late progression, and four of them had a Y3 sample: MRD was detected in three patients (lead time range 2.77-13.47 years), one patient had breast-only progression and was MRD-negative.

**Conclusions:** MRD status at key timepoints is associated with exceptional response and late distant progression, supporting prospective clinical trials implementing MRD testing with highly sensitive tumor-informed assays to guide treatment de-escalation.

## Introduction

Human epidermal growth factor receptor 2 (HER2)-positive (HER2+) tumors account for approximately 15% of all breast cancer diagnoses^1^. Following the development of HER2-targeted agents, most patients with early-stage disease are now cured^2–5^, and median overall survival (OS) in the metastatic setting exceeds five years^6^.

A subset of patients with HER2+ metastatic disease experiences exceptionally favorable outcomes, remaining on first-line therapy for years without evidence of disease progression. In the end-of-study analysis of the CLEOPATRA trial, 37% of patients treated with taxane, trastuzumab and pertuzumab (THP) were alive 8 years after starting treatment, and 16% had no evidence of disease progression^7^. Real-world studies have confirmed the extraordinary outcomes of some patients with HER2+ MBC^8, 9^. However, the paradigm of treatment for HER2+ MBC is palliative; though some retrospective series have shown excellent outcomes for patients who interrupt anti-HER2 therapies, prospective data supporting treatment cessation are lacking, and no predictive biomarkers are available to guide therapy in this setting^10, 11^. Clinicopathologic features like high membrane HER2 expression by immunohistochemistry (IHC), *de novo* metastatic disease presentation, oligometastatic disease, or radiologic complete response have been associated with prolonged benefit^12–16^, but none has been able to significantly stratify patients according to risk of future progression. How to select patients who could safely interrupt maintenance therapy is thus unknown.

The detection of Minimal Residual Disease (MRD) via circulating tumor DNA (ctDNA) is prognostic in patients with solid tumors, including breast cancer^17^. In the early-stage setting, virtually all patients with MRD detection after treatment experience distant recurrence^18–21^. Furthermore, the development of highly sensitive, tumor-informed MRD assays reduces the risk of false negative results and maximizes the lead-time between molecular and clinical relapse^19, 22–24^. Less evidence is available in the metastatic setting, where tracking MRD could provide treatment guidance for exceptional responders^25, 26^. No data are available with tumor-informed assays.

Here, we report the results of a case-control study investigating the association between MRD detection via the highly sensitive MAESTRO assay and response to HER2 targeted therapy by comparing the MRD status at key landmark timepoints between exceptional and conventional responders.

## Methods

### Patient Characteristics

We queried a prospective institutional database of patients with MBC seen at the Dana-Farber Cancer Institute and consented to the EMBRACE research cohort study (Dana-Farber/Harvard Cancer Center [DF/HCC] IRB #09-204), and identified all patients with HER2+ tumors by American Society of Clinical Oncology/ College of American Pathologists guidelines^27^ treated with first-line HER2 targeted therapy. Patients were diagnosed from 2008 to 2021, and the data cutoff was August 8, 2025. Patients with a real-world progression-free survival (rwPFS) equal to or longer than 3 years were considered exceptional responders; conventional responders had rwPFS shorter than 3 years. Within these two groups, we selected patients with archival tumor tissue, buffy coat, and at least one plasma sample collected at key landmark timepoints while on first-line therapy (e.g., at baseline, 1 year [Y1], 2 years [Y2], 3 years [Y3] after first-line therapy start). Baseline samples were collected after MBC diagnosis and before treatment start. Y1, Y2 and Y3 samples were identified considering a ±3-month window. For patients with any of the above key timepoints available, we also analyzed the first sample collected while on therapy (tx) (first-on-tx; up to 9 months after treatment start) and samples collected at time of progressive disease (PD) (within 30 days of progression), when available.

This study, which was conducted in accordance with the precepts established by the Declaration of Helsinki, was deemed exempt from IRB review (DF/HCC IRB #22-180) and granted access to archival clinical data and samples.

### Cell-free DNA (ctDNA) analysis

cfDNA was extracted from 0.3-6.6 mL archival plasma aliquots, as previously described^28^. When plasma was not available, previously extracted cfDNA or cfDNA libraries were used instead (**Supplementary Table 1**). Whole-genome sequencing (WGS) was carried out on archival tumor tissue collected at any timepoint (i.e., primary or metastasis) and matched normal genomic DNA from buffy coat samples to identify somatic mutations. These somatic mutations were used to design MAESTRO mutation enrichment sequencing assays, as previously described^28^. MAESTRO assays were then applied to all patient-specific cfDNA, tumor tissue and matched normal samples in a pooled format that applies all bespoke assays to all samples from all patients (MAESTRO-Pool), as previously described^29, 30^. After consensus calling and filtering, we retained only the mutations that were validated in the tumor tissue and not detected in the matched normal to ensure true somatic mutations were tracked. Finally, MRD status, tumor fraction (TFx) and limit of detection 95% (LOD95) were generated^30^. TFx was defined as the fraction of tumor-derived cfDNA fragments; LOD95 as the minimal TFx estimated to be detected with 95% probability based on the number of cfDNA molecules and tumor mutations assayed in each sample.

### Study Design and Statistical Analysis

The primary objective of this case-control study was to investigate the association between MRD status at the Y1 timepoint and exceptional response. Since Y1 samples were collected with a ±3 months window, we considered a landmark timepoint of 15 months for the analysis; patients who experienced disease progression within 15 months of treatment start were excluded. In exploratory analyses we compared clinicopathologic features between exceptional and conventional responders and described the evolution of MRD over time and the distribution of TFx at various timepoints.

For the primary analysis, rwPFS was defined from the start of first-line therapy to the first progression event that led to treatment switch or death, whichever occurred first. We prospectively chose to include only events that led to treatment switch to exclude local and brain-only progression events, which are usually treated with locoregional therapy only and often not detected by MRD assays. In sensitivity analyses, we used alternative definitions of rwPFS that considered: i) all progression events, including local and brain-only progression, regardless of treatment switch, or ii) progression events that led to treatment switch but excluded death. Disease progression was adjudicated by the investigators based on review of the medical record. The 3-year benchmark for rwPFS was chosen following the National Cancer Institute definition of Exceptional Response^31^ considering duration of benefit exceeding three times the median PFS in the control arm of CLEOPATRA (e.g., 12 months)^32^. rwPFS and OS were analyzed via the Kaplan-Meier method and with a landmark analysis at year 3 for exceptional responders. Median and rate of survival were reported with 95% confidence interval (CI). Fisher exact test was used to assess the association between response groups and MRD status, with significance defined to be at p<0.05.

## Results

### Patients Characteristics and Survival Outcomes

We identified 70 patients with HER2+ MBC treated with first-line HER2 targeted therapy and available archival tumor tissue, buffy coat and plasma for at least one of the key time points. Sixty-three patients (90%) - 40 exceptional and 23 conventional responders - had sufficient samples and successful assay design and were included in the final analysis (**Figure 1**).

**Figure 1:**
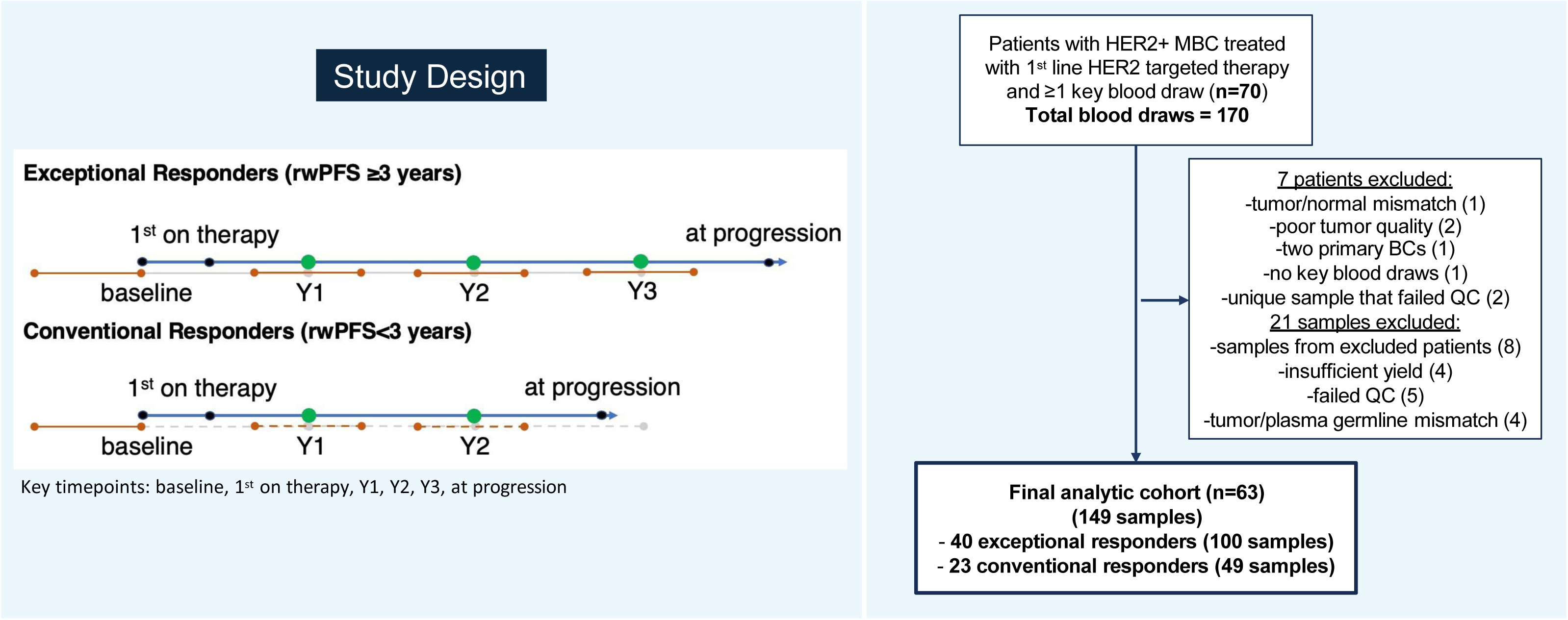
Study Design and REMARK Diagram. **Abbreviations:** BCs, breast cancers; HER2, human epidermal growth factor receptor 2; QC; quality check; MBC, metastatic breast cancer; rwPFS, real-world progression-free survival; Y1, year 1; Y2, year 2; Y3, year 3. Timing of sample collection for each timepoint is defined in Table 2.

Patient and tumor characteristics are compared between exceptional and conventional responders in **Table 1**. All features were similar between the two groups, except for the proportion of patients with *de novo* metastatic disease that was higher among exceptional than conventional responders though not significantly different (70% vs. 43.5%, p=0.061). Among both exceptional and conventional responders, most patients had high (3+ by IHC) HER2 expression (85% vs. 73.9%), ER-negative disease (60% vs 65.2%), and visceral metastases at diagnosis (87.5% vs. 65.2%). The most common first-line regimen was chemotherapy plus trastuzumab/pertuzumab in both groups (82.5% vs. 95.7%), and most patients received maintenance endocrine therapy if estrogen receptor (ER)-positive (93.3% vs. 62.5%).

**Table 1:** Clinicopathological Features of Exceptional and Conventional Responders.

|  | **Conventional Responders (N=23)** | **Exceptional Responders (N=40)** | **Overall Population (N=63)** | **p value** |
| --- | --- | --- | --- | --- |
| **Age at Metastatic Diagnosis** |  |  |  | 0.392 |
| Median | 50.3 | 48.0 | 48.5 |  |
| Range | 26.9 - 72.8 | 26.0 - 72.3 | 26.0 - 72.8 |  |
| **Racial Background** |  |  |  | 0.238 |
| African American | 4 (17.4%) | 1 (2.5%) | 5 (7.9%) |  |
| American Indian, Aleutian, Eskimo | 0 (0.0%) | 1 (2.5%) | 1 (1.6%) |  |
| Asian or Pacific Islander | 1 (4.3%) | 3 (7.5%) | 4 (6.3%) |  |
| White | 18 (78.3%) | 34 (85.0%) | 52 (82.5%) |  |
| Other | 0 (0.0%) | 1 (2.5%) | 1 (1.6%) |  |
| **Tumor Stage at Diagnosis** |  |  |  | 0.061 |
| I, II, III | 13 (56.5%) | 12 (30.0%) | 25 (39.7%) |  |
| IV | 10 (43.5%) | 28 (70.0%) | 38 (60.3%) |  |
| **HER2 Status** |  |  |  | 0.328 |
| 0-2+/FISH-positive | 6 (26.1%) | 6 (15.0%) | 12 (19.0%) |  |
| 3+ | 17 (73.9%) | 34 (85.0%) | 51 (81.0%) |  |
| **Estrogen Receptor (ER)** |  |  |  | 0.790 |
| Positive | 8 (34.8%) | 16 (40.0%) | 24 (38.1%) |  |
| Negative | 15 (65.2%) | 24 (60.0%) | 39 (61.9%) |  |
| **Visceral Disease** |  |  |  | 0.546 |
| Yes | 19 (82.6%) | 30 (75.0%) | 49 (77.8%) |  |
| No | 4 (17.4%) | 10 (25.0%) | 14 (22.2%) |  |
| **Brain Metastases** |  |  |  | 0.511 |
| Yes | 5 (21.7%) | 6 (15.0%) | 11 (17.5%) |  |
| No | 18 (78.3%) | 34 (85.0%) | 52 (82.5%) |  |
| **First Line Regimen\*** |  |  |  | 0.522 |
| Chemotherapy (non-taxane) plus trastuzumab | 2 (8.7%) | 1 (2.5%) | 3 (4.8%) |  |
| Chemotherapy (non-taxane) plus trastuzumab/pertuzumab | 3 (13.0%) | 0 (0.0%) | 3 (4.8%) |  |
| Chemotherapy (taxane) plus trastuzumab/pertuzumab | 14 (60.9%) | 33 (82.5%) | 47 (74.6%) |  |
| Lapatinib/trastuzumab | 0 (0.0%) | 1 (2.5%) | 1 (1.6%) |  |
| T-DM1/neratinib | 0 (0.0%) | 1 (2.5%) | 1 (1.6%) |  |
| T-DM1 | 3 (13.0%) | 0 (0.0%) | 3 (4.8%) |  |
| Trastuzumab (+/- endocrine therapy) | 1 (4.3%) | 0 (0.0%) | 1 (1.6%) |  |
| Trastuzumab/pertuzumab (+/- endocrine therapy) | 0 (0.0%) | 4 (10.0%) | 4 (6.3%) |  |
| **Maintenance Endocrine therapy (if ER-Positive)** |  |  |  | 0.289 |
| Yes | 5 (62.5%) | 14 (87.5%) | 19 (79.2%) |  |
| No | 3 (37.5%) | 2 (12.5%) | 5 (20.8%) |  |
| *The p value was calculated comparing chemotherapy plus trastuzumab/pertuzumab (taxane and non-taxane based) and the rest of the categories. |  |  |  |  |

**Abbreviations:** HER2, human epidermal growth factor receptor 2; FISH, Fluorescence In Situ Hybridization; T-DM1, ado-trastuzumab emtansine

With 289.9 person-years of follow up, six exceptional responders experienced late progression (e.g., after year 3). Since exceptional responders were progression-free for at least three years by the study criteria, survival outcomes were estimated by landmark analysis starting at year 3. Two-year rwPFS (e.g., 5 years from treatment start) was 94.9% (95% CI 81.0%, 98.7%), and 5-year OS (e.g., 8 years from treatment start) was 88.7% (95%CI 67.6%, 96.4%) (**Figure S1**). For conventional responders, total follow up was 101.3 person-years. Median rwPFS was 1.60 years (95%CI 1.1-2.51), and median OS was 4.58 yrs (95%CI 3.78-6.48) (**Figure S2**).

### MRD detection and quantification

Overall, a median of 1,823 (range 387-5,000) tumor-specific mutations were tracked per patient and MAESTRO-Pool was run on 149 samples (**Table S1**). MRD was detected in 49 (32.9%) samples, including 6/7 (85.7%) baseline, 14/43 (32.6%) first-on-tx, 14/44 (31.8%) Y1, 5/30 (16.7%) Y2, 3/18 (16.7%) Y3, 7/7 (100%) at progression (**Table 2**). Median TFx across all MRD-positive samples was 936 ppm (range 3.8-164,068 ppm) (**Figure 2**). Median LOD95 (i.e., the lowest TFx detectable with 95% likelihood) across all samples was 6.45 ppm (IQR 4.43-13.55 ppm), and 6.43 ppm across the 100 MRD negative samples (IQR 4.48-13.26 ppm) (**Table S1**). Four samples were considered underpowered, as the LOD95 was greater than 100 ppm. Since each patient’s bespoke MRD test was applied to all plasma samples from all patients in the cohort using MAESTRO-Pool, we also computed an overall specificity for ctDNA detection from all “patient-unmatched” MRD tests. Among 9238 patient-unmatched MRD tests, only 26 were detected (median tumor fraction 4ppm, range 2ppm-55ppm), yielding an overall specificity of 99.72%.

**Figure 2:**
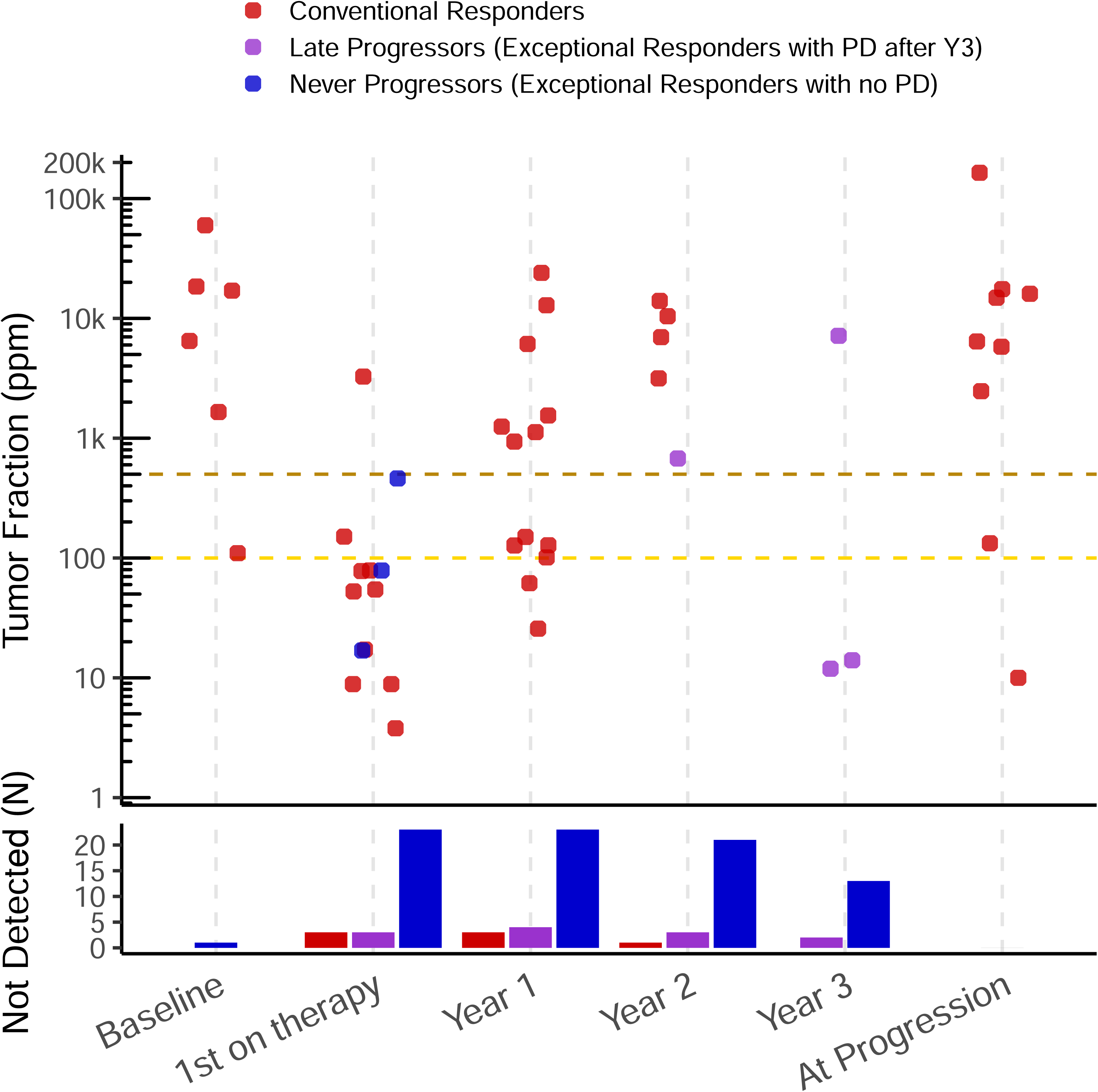
Distribution of TFx at various timepoints, by response group. **Abbreviations:** PD, progressive disease; TFx, tumor fraction; Y3, year 3

**Table 2:** MRD status at key timepoints by response group.

| **Timepoint** | **Conventional Responders (49 samples)** | **Exceptional Responders (100 samples)** |
| --- | --- | --- |
| Baseline<sup>1</sup> | 6/6 (100%) | 0/1 (0%) |
| First-on-therapy (First-on-tx)<sup>2</sup> | 11/14 (79%) | 3/29 (10%) |
| Year 1 (Y1)<sup>3</sup> | 14/17 (82%) | 0/27 (0%) |
| Year 2 (Y2)<sup>4</sup> | 4/5 (80%) | 1/25 (4%) |
| Year 3 (Y3)<sup>5</sup> | Not Applicable | 3/18 (17%) |
| At time of disease progression (at PD)<sup>6</sup> | 7/7 (100%) | Not Available |
| <sup>1</sup>Before 1L treatment start<br><sup>2</sup>First sample collected on treatment, up to 9 months after 1L treatment start<br><sup>3</sup>281-449 days after 1L treatment start<br><sup>4</sup>646-814 days after 1L treatment start<br><sup>5</sup>1011-1179 days after 1L treatment start<br><sup>6</sup>Within 30 days prior to disease progression leading to treatment switch |  |  |

**Abbreviations: MRD, minimal residual disease; Y, year; PD, progressive disease; 1L, first-line; tx, therapy**

In an exploratory analysis, we simulated how the detection rate would have changed if our assay had been less sensitive, considering 500 ppm (e.g., 0.05%) and 100 ppm (e.g., 0.01%), LOD95 thresholds as per many commercially available tests in the early-stage and metastatic setting, respectively. At 500 ppm, we would not have detected MRD in 23/49 (46.9%) samples, including 1/6 baseline, 12/14 first-on-tx, 6/14 Y1, 2/3 Y3, 2/7 at progression. At 100 ppm, we would not have detected MRD in 15/49 (30.6%) samples, including 10/14 first-on-tx, 2/14 Y1, 2/3 Y3, 1/7 at PD (**Figure 2**). Our findings suggest that highly sensitive assays are better suited to identify true MRD-negative cases.

### MRD status and Exceptional Response to HER2 targeted therapy

Next, we sought to investigate the association between MRD status and exceptional response. After excluding samples collected at baseline and at progression, which are expected to be MRD-positive, MRD was detected in 7/99 (7.1%) and 29/36 (80.6%) samples from exceptional and conventional responders, respectively. All exceptional responders who remained progression-free were always MRD-negative (n=30) or cleared MRD by Y1 (n=3) (**Figure 3**). Indeed, Y1 MRD status was associated with exceptional versus conventional response (0/27 [0%] versus 9/12 [75.0%] had detectable MRD, p<0.001). No baseline clinicopathological features were associated with Y1 MRD status (**Table S2**), suggesting the role of MRD as an independent factor associated with exceptional response. Similar findings were observed when investigating the association between first-on-tx MRD status (collected at a median of 4 months [range 0.23-9.2] after first-line therapy start) and exceptional response: 3/29 (10.3%) versus 10/13 (76.9%) samples were MRD-positive in exceptional and conventional responders, respectively (p<0.001). The three MRD-positive samples from exceptional responders were collected 0.22-1.6 months from treatment start, suggesting that ctDNA clearance might not have occurred by the time of sampling.

**Figure 3:**
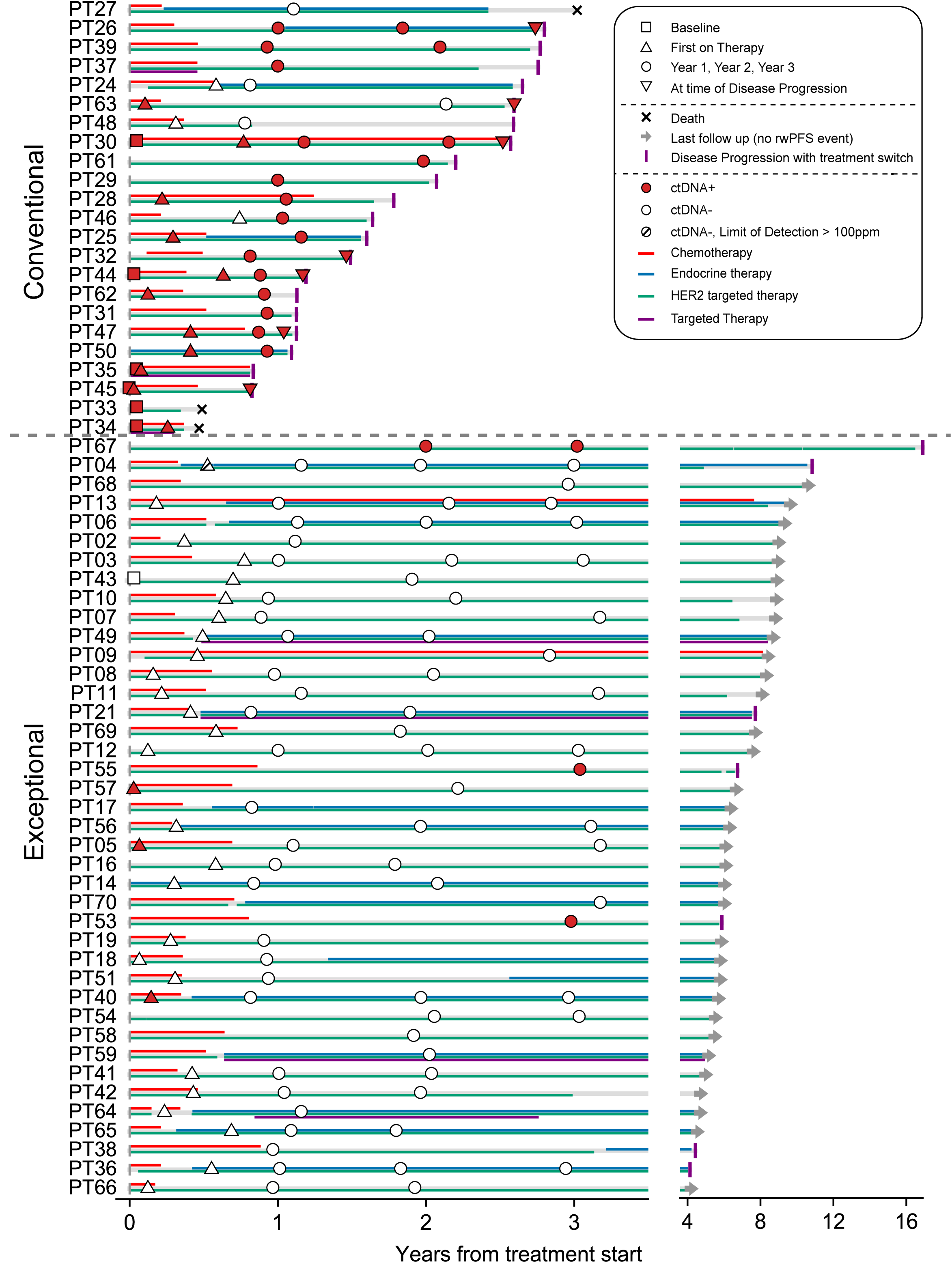
Study Cohort, swimmers plot. **Abbreviations:** ctDNA, circulating tumor DNA; HER2, human epidermal growth factor receptor 2.

To investigate whether Y3 MRD status was associated with late progression (e.g., after year 3), we looked at progression events among exceptional responders with a Y3 sample available. Of the seven exceptional responders with late disease progression (patients 04, 21, 36, 38, 53, 55, 67; **Figure 3**), five patients had a Y3 draw (patients 04, 36, 53, 55, 67). Three of the five samples were MRD-positive, with a lead time between collection and progression of 2.74, 3.59 and13.48 years. Patient 04 who had Y3 MRD-negative sample experienced breast-only progression, whereas MRD was not detected at Y3 in patient 36 despite subsequent disease progression. Hence, our findings suggest that MRD status at Y3 is predictive of late distant progression among most exceptional responders.

Given the limited number of patients with multiple samples, no formal analyses of TFx dynamics were conducted (**Figure S3**).

Finally, we investigated why five conventional responders had some MRD-negative samples (3/13 first-on-tx, 3/16 Y1, 1/5 Y2).. For all patients, samples were collected at time of complete response (patients 46, 63) and/or minimal tumor burden (patients 24, 48, 27). Furthermore, some of these samples consisted of previously extracted cfDNA (patient 24; LOD95 13.59 ppm for first-on-tx and 98.22ppm for Y1 sample), potentially compromising specimen quality, or were from patients with a small panel size (patient 46, tracking 469 sites, LOD95 42.81 ppm for first-on-tx sample).

### Sensitivity Analyses with different rwPFS definitions

To investigate whether our primary definition of rwPFS was too strict, we conducted a sensitivity analysis including all disease progressions or deaths as events, regardless of treatment switch. With this definition, six exceptional responders were reclassified as conventional responders, and 10 patients remained in the same category but experienced disease progression earlier than in the original definition. The primary endpoint of association between MRD status at Y1 and response remained significant (p<0.001). Still, no exceptional responders were MRD-positive at Y1 (0/23, 0%) but the proportion of conventional responders positive at Y1 decreased (8/14, 57.1%). 11/16 early progression events occurred in the brain, and six were recorded among exceptional responders who experienced progression that was treated locally without systemic treatment switch. This study was not designed to assess the role of MRD in predicting brain-only progression, so samples collected at time of limited brain progression treated with local therapy were not included. However, two patients (25 and 47) had brain-only progression that led to treatment switch, and both had MRD+ samples prior to or at time of progression, suggesting that significant brain progression can be detected by MRD assays.

Conversely, a stricter definition of rwPFS that included only progression events that led to treatment switch and excluded deaths led to more significant results: no exceptional responders were positive at Y1 (0/27, 0%), compared to nine of 11 conventional responders (81.8%; p<0.001).

## Discussion

A significant subset of patients with HER2+ MBC experiences exceptional response and remains on first-line therapy without progression for many years. However, whether some patients can safely interrupt or permanently stop therapy is unknown, in absence of an understanding of the biology of exceptional responses and predictive biomarkers to guide therapy. Here, we report results of a case-control study investigating the role of ultrasensitive MRD testing to predict exceptional response to first-line HER2 targeted therapy for MBC. MAESTRO-Pool was run on 149 samples collected at key timepoints from 40 exceptional responders and 23 conventional responders. In our study, MRD status at Y1 was significantly associated with exceptional response, as all exceptional responders who remained progression-free were always MRD negative or cleared MRD by Y1; 75% of conventional responders were MRD-positive at Y1. Furthermore, MRD status at Y3 was associated with late distant progression, though this time point was available for only a small group of patients. We also observed that ∼30% of MRD-positive samples using the MAESTRO-Pool assay had TFx below the LOD of first-generation, tumor-informed MRD assays.

To our knowledge, this is the largest study investigating MRD in exceptional responders with MBC using a highly sensitive assay. For comparison, PRE-PHENIX included 40 exceptional responders and 11 controls and tested MRD with a tumor agnostic assay with a limit of quantification of 0.01%. MRD was detected in 10% of long-term responders who were on maintenance therapy for at least 4 years, and 91% of conventional responders at time of confirmed disease progression^26^. No data were available on late distant progression among long-term responders to discriminate whether the positive samples were false positive or associated with subsequent progression. In preliminary data from the HEROES study, prevalence of MRD was 8% among exceptional responders who were progression-free for at least 2 years using a whole exome sequencing (WES)-based, tumor-informed assay, but no data on late progression events are available^33^.

Our results suggest that MRD status at Y1 is a strong predictor of exceptional response. Though not all conventional responders were MRD-positive at every timepoint, an MRD-positive status was significantly associated with progression before reaching the 3-year timepoint. This is relevant in the setting of the evolving therapeutic landscape of first-line therapy for HER2+ MBC. The CLEOPATRA regimen of THP induction followed by indefinite HP maintenance has been the recommended first-line regimen for more than a decade, but the combination of Trastuzumab Deruxtecan (T-DXd) plus pertuzumab almost doubled PFS compared to THP in the DESTINY-Breast09 trial (median PFS 40.7 versus 26.9 months, p<0.00001), and is likely to become the recommended first-line option^34^. Furthermore, in the PATINA and HER2CLIMB-05 maintenance studies adding respectively palbociclib to HP and endocrine therapy for HER2+/ER+ tumors or tucatinib to maintenance HP regardless of ER expression significantly delayed the risk of progression^35, 36^. In the absence of head-to-head comparisons, how to best choose therapeutic options is unknown. Our data suggest that MRD status at Y1 might help to select patients who would benefit from treatment de-escalation. Furthermore, absence of MRD clearance might help select those who could benefit from treatment escalation or switch. Prospective studies are required to test these hypotheses.

Our primary definition of rwPFS excluded brain- and local-only progression events as those are usually treated with locoregional therapy and prior studies showed a limited sensitivity of MRD tests for these events. However, to confirm that this choice did not significantly bias the results, we conducted a sensitivity analysis considering all progression events, and MRD status at Y1 remained a strong predictor of exceptional response. Six of the 11 early, brain-only events occurred in exceptional responders who never subsequently experienced systemic progression, suggesting that brain metastases do not necessarily compromise the possibility of achieving exceptional response. No samples were analyzed at time of brain-only progression that was treated locoregionally, so it is unknown whether MRD was detectable at that time. However, samples collected prior to those events were MRD-negative. Conversely, MRD was detected at the time of more significant brain-only progression that was treated by switching systemic therapy, suggesting that multifocal and/or large brain progression might be detected by highly sensitive assays.

In our study, MRD status at Y3 was associated with late distant progression. How to best treat exceptional responders is unknown, and multiple studies are investigating whether HER2 therapy can be safely stopped. In our fully accrued STOPHER2 trial (NCT05721248, TBCRC-062), patients on first-line therapy for at least three years without evidence of disease progression chose whether to stop or continue treatment, while undergoing close imaging surveillance and providing plasma samples for retrospective MRD analysis. We expect to report the primary endpoint – 1-year PFS off therapy - in 2026. The HEROES study (NCT06450314) is instead investigating treatment interruption only in patients with disease controlled after 2 years of anti-HER2 maintenance therapy who are MRD-negative with a first-generation, WES-based tumor-informed assay^33^. Finally, Free-HER (NCT05959291) is a pilot study enrolling 20 patients in complete response for at least 3 years and negative ctDNA results with a tumor-informed, WES-based assay or, in absence of adequate tissue, with a tumor-agnostic assay^37^. In our study, all patients who were MRD-positive at Y3 experienced late distant progression, suggesting that MRD at this timepoint might help guide treatment discontinuation. Of note, two of the three MRD-positive samples at Y3 had TFx<100ppm, underscoring the importance of using highly sensitive tests to avoid false negative results.

Our study has some limitations. This is a retrospective, single center, real-world study. Intervals between restaging scans were heterogeneous, and blood samples were not available at all time points for all patients. Treatments were also heterogeneous; racial and ethnic diversity was limited. Because our cohort study does not collect progression data using RECIST criteria, we used rwPFS. Few baseline samples were available so we could not assess the rate and timing of MRD clearance. Furthermore, some samples had limited plasma volume and for others only previously extracted cfDNA was available. Finally, the sample size was relatively small for the overall population and for each timepoint, limiting the power of the study.

In conclusion, HER2+ MBC has been considered an incurable disease, but the availability of highly effective targeted therapies is challenging this dogma. Studies like STOP-HER2, HEROES and Free-HER are investigating if maintenance therapy can be safely stopped. In parallel, the SAPPHO trial (NCT06439693, TBCRC-065) is investigating whether treatment intensification through sequential, non-cross resistant HER2-targeted regimens followed by treatment cessation can increase the proportion of exceptional responders. In routine practice, the use of first-line T-DXd plus pertuzumab until progression or toxicity, as established by the DESTINY-Breast09 trial, will likely lead to a higher proportion of patients with exceptional response^34^. However, how to optimally tailor HER2-targeted therapies is unknown, and studies investigating dynamics of MRD to guide treatment are needed. Our findings suggest that MRD may be a powerful tool to predict exceptional response early, but that highly sensitive assays are needed to minimize the risk of false-negative MRD results. Prospective studies investigating how to best use MRD to guide therapy for HER2+ MBC are warranted.

## Data Availability

The data generated in this study are available within the article and
its Supplementary Data files. Additional data may be provided upon request.

## Acknowledgements

The authors greatly appreciate the support of Valerie Hope Goldstein (editor, full-time employee of Dana-Farber Cancer Institute) in formatting and submitting this manuscript.

## Data Sharing Statement

The data generated in this study are available within the article and its Supplementary Data files. Additional data may be provided upon request.

## Author Contributions

**Conceptualization:** S.M., N.U.L., V.A.A., H.A.P.

**Methodology:** S.M., C.So., N.Z., H.H., N.T., V.A.A., H.A.P.

**Software:** S.M., C.So., N.Z., J.R., G.K., H.H., V.A.A.

**Validation:** C.So., P.J., L.W., R.L., J.R.

**Formal analysis:** S.M., C.So., N.Z., H.H.

**Investigation:** S.M., K.S., P.J., L.W., R.L.

**Resources:** S.T., N.U.L., V.A.A., H.A.P.

**Data Curation:** S.M., C.So., N.Z., K.S., C.St., J.R., G.K.

**Writing - Original Draft:** S.M., C.So.

**Writing - Review & Editing:** All authors

**Visualization:** S.M., C.So.

**Supervision:** J.R., A.P., M.E.H., N.T., H.H., K.X., N.U.L., V.A.A., H.A.P

**Project administration:** K.G., K.X., V.A.A., H.A.P

**Funding acquisition:** S.M., G.C., N.U.L., V.A.A., H.A.P

## References

1. Giaquinto AN, Sung H, Newman LA, et al: Breast cancer statistics 2024 [Internet]. CA Cancer J Clin, 2024[cited 2024 Oct 4] Available from: https://onlinelibrary.wiley.com/doi/abs/10.3322/caac.21863

2. . Harbeck N, Modi S, Pusztai L, et al: Neoadjuvant trastuzumab deruxtecan alone or followed by paclitaxel, trastuzumab, and pertuzumab for high-risk HER2-positive early breast cancer (DESTINY-Breast11): a randomised, open-label, multicentre, phase 3 trial [Internet]. Ann Oncol 0, 2025[cited 2025 Nov 19] Available from: 10.1016/j.annonc.2025.10.019

3. . von Minckwitz G, Huang C-S, Mano MS, et al: Trastuzumab Emtansine for Residual Invasive HER2-Positive Breast Cancer. N Engl J Med 380:617–628, 2019

4. . von Minckwitz Gunter, Procter Marion, de Azambuja Evandro, et al: Adjuvant Pertuzumab and Trastuzumab in Early HER2-Positive Breast Cancer. N Engl J Med 377:122–131

5. . Tolaney SM, Tarantino P, Graham N, et al: Adjuvant paclitaxel and trastuzumab for node-negative, HER2-positive breast cancer: final 10-year analysis of the open-label, single-arm, phase 2 APT trial. Lancet Oncol 24:273–285, 2023

6. . Grinda T, Antoine A, Jacot W, et al: Evolution of overall survival and receipt of new therapies by subtype among 20 446 metastatic breast cancer patients in the 2008-2017 ESME cohort. ESMO Open 6:100114, 2021

7. . Swain SM, Miles D, Kim SB, et al: Pertuzumab, trastuzumab, and docetaxel for HER2-positive metastatic breast cancer (CLEOPATRA): end-of-study results from a double-blind, randomised, placebo-controlled, phase 3 study. Lancet Oncol 21:519–530, 2020

8. . Wong Y, Raghavendra AS, Hatzis C, et al: Long-Term Survival of De Novo Stage IV Human Epidermal Growth Receptor 2 (HER2) Positive Breast Cancers Treated with HER2-Targeted Therapy. Oncologist 24:313–318, 2019

9. . Gobbini E, Ezzalfani M, Dieras V, et al: Time trends of overall survival among metastatic breast cancer patients in the real-life ESME cohort. Eur J Cancer 96:17–24, 2018

10. Moilanen T, Mustanoja S, Karihtala P, et al: Retrospective analysis of HER2 therapy interruption in patients responding to the treatment in metastatic HER2+ breast cancer [Internet]. ESMO Open 2, 2017Available from: 10.1136/esmoopen-2017-000202

11. . Witzel I, Müller V, Abenhardt W, et al: Long-term tumor remission under trastuzumab treatment for HER2 positive metastatic breast cancer - results from the HER-OS patient registry [Internet]. BMC Cancer 14, 2014Available from: 10.1186/1471-2407-14-806

12. Debien V, Agostinetto E, Bruzzone M, et al: The Impact of Initial Tumor Response on Survival Outcomes of Patients With HER2-Positive Advanced Breast Cancer Treated With Docetaxel, Trastuzumab, and Pertuzumab: An Exploratory Analysis of the CLEOPATRA Trial [Internet]. Clin Breast Cancer, 2024Available from: https://www.sciencedirect.com/science/article/pii/S1526820924000508

13. . Veitch Z, Ribnikar D, Tilley D, et al: No evidence of disease versus residual disease in long-term responders to first-line HER2-targeted therapy for metastatic breast cancer. Br J Cancer 126:881–888, 2022

14. . Murthy P, Kidwell KM, Schott AF, et al: Clinical predictors of long-term survival in HER2-positive metastatic breast cancer. Breast Cancer Res Treat 155:589–595, 2016

15. . Omarini C, Bettelli S, Caprera C, et al: Clinical and molecular predictors of long-term response in HER2 positive metastatic breast cancer patients. Cancer Biol Ther 19:879–886, 2018

16. . Yardley DA, Tripathy D, Brufsky AM, et al: Long-term survivor characteristics in HER2-positive metastatic breast cancer from registHER. Br J Cancer 110:2756–2764, 2014

17. . Moding EJ, Nabet BY, Alizadeh AA, et al: Detecting Liquid Remnants of Solid Tumors: Circulating Tumor DNA Minimal Residual Disease. Cancer Discov 11:2968–2986, 2021

18. . Lipsyc-Sharf M, de Bruin EC, Santos K, et al: Circulating Tumor DNA and Late Recurrence in High-Risk Hormone Receptor-Positive, Human Epidermal Growth Factor Receptor 2-Negative Breast Cancer. J Clin Oncol 15:2408–2419, 2022

19. Garcia-Murillas I, Abbott CW, Cutts RJ, et al: Whole genome sequencing powered ctDNA sequencing for breast cancer detection [Internet]. Ann Oncol 0, 2025[cited 2025 Feb 7] Available from: http://www.annalsofoncology.org/article/S0923753425000535/abstract

20. . Shaw JA, Page K, Wren E, et al: Serial Postoperative Circulating Tumor DNA Assessment Has Strong Prognostic Value During Long-Term Follow-Up in Patients With Breast Cancer. JCO Precis Oncol 8:e2300456, 2024

21. . Parsons HA, Rhoades J, Reed SC, et al: Sensitive Detection of Minimal Residual Disease in Patients Treated for Early-Stage Breast Cancer. Clin Cancer Res 26:2556–2564, 2020

22. . Coakley M, Villacampa G, Sritharan P, et al: Comparison of circulating tumor DNA assays for molecular residual disease detection in early-stage triple-negative breast cancer. Clin Cancer Res 30:895–903, 2024

23. . Parsons HA, Blewett T, Chu X, et al: Circulating tumor DNA association with residual cancer burden after neoadjuvant chemotherapy in triple-negative breast cancer in TBCRC 030☆. Ann Oncol 34:899–906, 2023

24. . Patel AK, Rhoades J, Xiong K, et al: 404P MRD analysis following curative intent treatment in patients with locally advanced esophageal cancer utilizing MAESTRO, an ultrasensitive ctDNA assay. Ann Oncol 36:S158, 2025

25. . Hunter N, Croessmann S, Cravero K, et al: Undetectable tumor cell-free DNA in a patient with metastatic breast cancer with complete response and long-term remission. J Natl Compr Canc Netw 18:375–379, 2020

26. . Llombart-Cussac A, Fernandez-Murga L, Martinez E, et al: Circulating tumor DNA (ctDNA)-based minimal residual disease (MRD) measured by Guardant Reveal in patients (pts) with HER2-positive (HER2+) metastatic breast cancer (mBC) with long-term disease control on first-line trastuzumab-pertuzumab. J Clin Oncol 43:1052–1052, 2025

27. . Wolff AC, Somerfield MR, Dowsett M, et al: Human epidermal growth factor receptor 2 testing in breast cancer: ASCO-College of American Pathologists guideline update. J Clin Oncol 41:3867–3872, 2023

28. . Adalsteinsson VA, Ha G, Freeman SS, et al: Scalable whole-exome sequencing of cell-free DNA reveals high concordance with metastatic tumors. Nat Commun 8:1–13, 2017

29. . Gydush G, Nguyen E, Bae JH, et al: Massively parallel enrichment of low-frequency alleles enables duplex sequencing at low depth. Nat Biomed Eng 6:257–266, 2022

30. . Blewett T, Rhoades J, Liu R, et al: MAESTRO-Pool enables highly parallel and specific mutation-enrichment sequencing for minimal residual disease detection in cohort studies. Clin Chem 70:434–443, 2024

31. . Wheeler DA, Takebe N, Hinoue T, et al: Molecular Features of Cancers Exhibiting Exceptional Responses to Treatment. Cancer Cell 39:38–53.e7, 2021

32. . Baselga J, Cortés J, Kim S-B, et al: Pertuzumab plus trastuzumab plus docetaxel for metastatic breast cancer. N Engl J Med 366:109–119, 2012

33. De La Motte Rouge T, Michiels S, Hardy-Bessard A-C, et al: Heroes: De-escalation of medical therapies in her2-positive metastatic breast cancer in longterm persistent response and minimal residual disease undetectable in circulating tumor dna

34. . Tolaney SM, Jiang Z, Zhang Q, et al: Trastuzumab deruxtecan plus pertuzumab for HER2-positive metastatic breast cancer [Internet]. N Engl J Med, 2025Available from: 10.1056/nejmoa2508668

35. . Metzger O, Mandrekar S, Goel S, et al: Palbociclib for hormone-receptor-positive, HER2-positive advanced breast cancer. N Engl J Med 394:451–462, 2026

36. . Dieras V, Curigliano G, Martin M, et al: HER2CLIMB-05: A phase 3 study of tucatinib versus placebo in combination with trastuzumab and pertuzumab as first-line maintenance therapy for HER2+ metastatic breast cancer. J Clin Oncol 101200JCO2502600, 2025

37. . Jackson EK, F. valdes, Calfa C, et al: Abstract PS1-10-20: Free-her: discontinuation of maintenance her-2 directed therapy in long-term survivors of metastatic her-2 positive breast cancer: a trial update. Clin Cancer Res 32:PS1–10-20-PS1-10–20, 2026

