## Supplement for "Minimal Residual Disease via circulating tumor DNA Predicts Exceptional Response in HER2-Positive Metastatic Breast Cancer"

**Figure S1:** Real-World Progression-Free Survival (rwPFS, top) and Overall Survival (OS, bottom) for Exceptional Responders

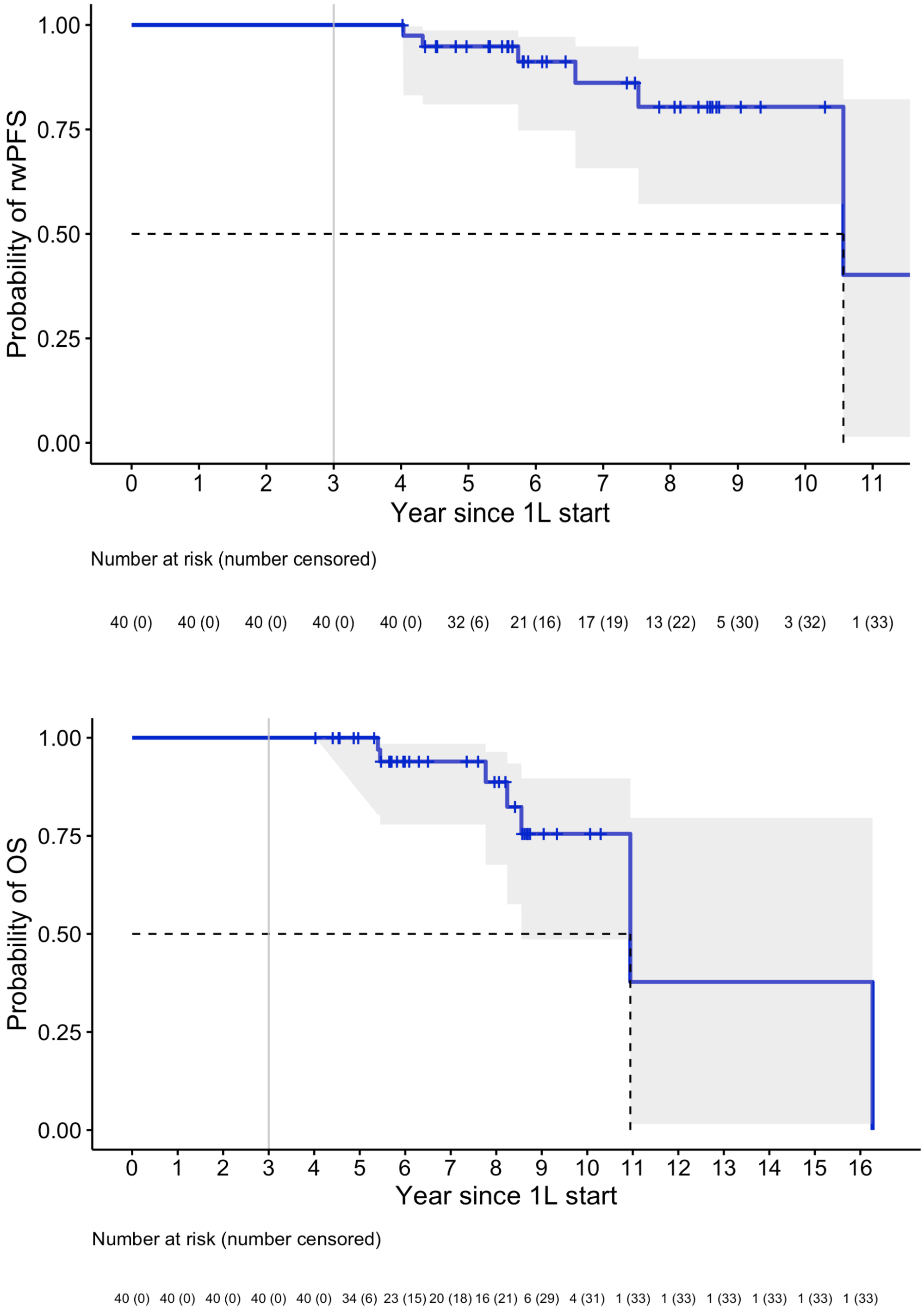

Abbreviations: 1L, first-line treatment

**Figure S2:** Real-World Progression-Free Survival (rwPFS, top) and Overall Survival (OS, bottom) for Conventional Responders.

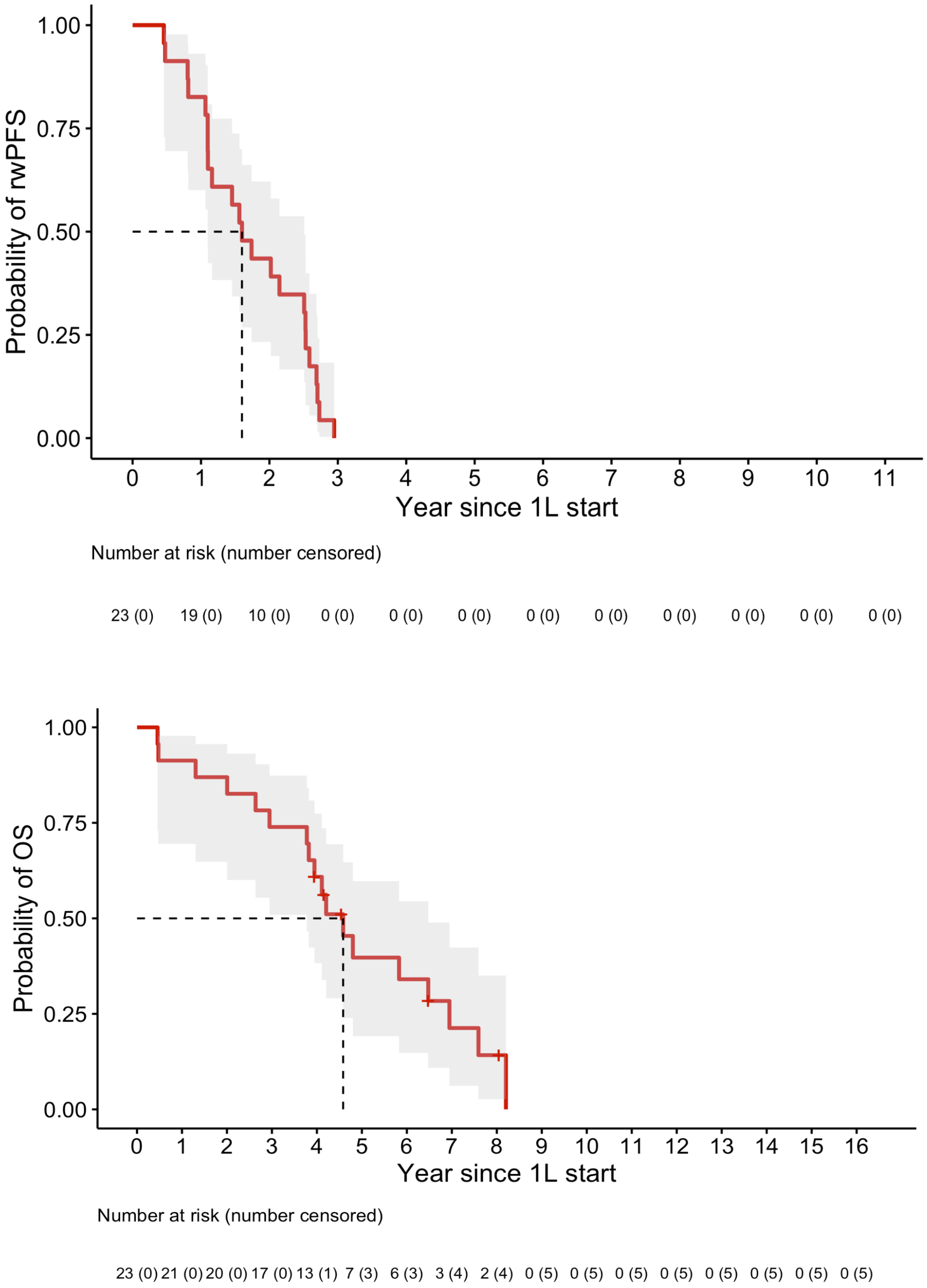

Abbreviations: 1L, first-line treatment

**Figure S3:** Patient-level, tumor fraction dynamics for patients with multiple timepoints available, separately for exceptional responders (top) and conventional responders (bottom).

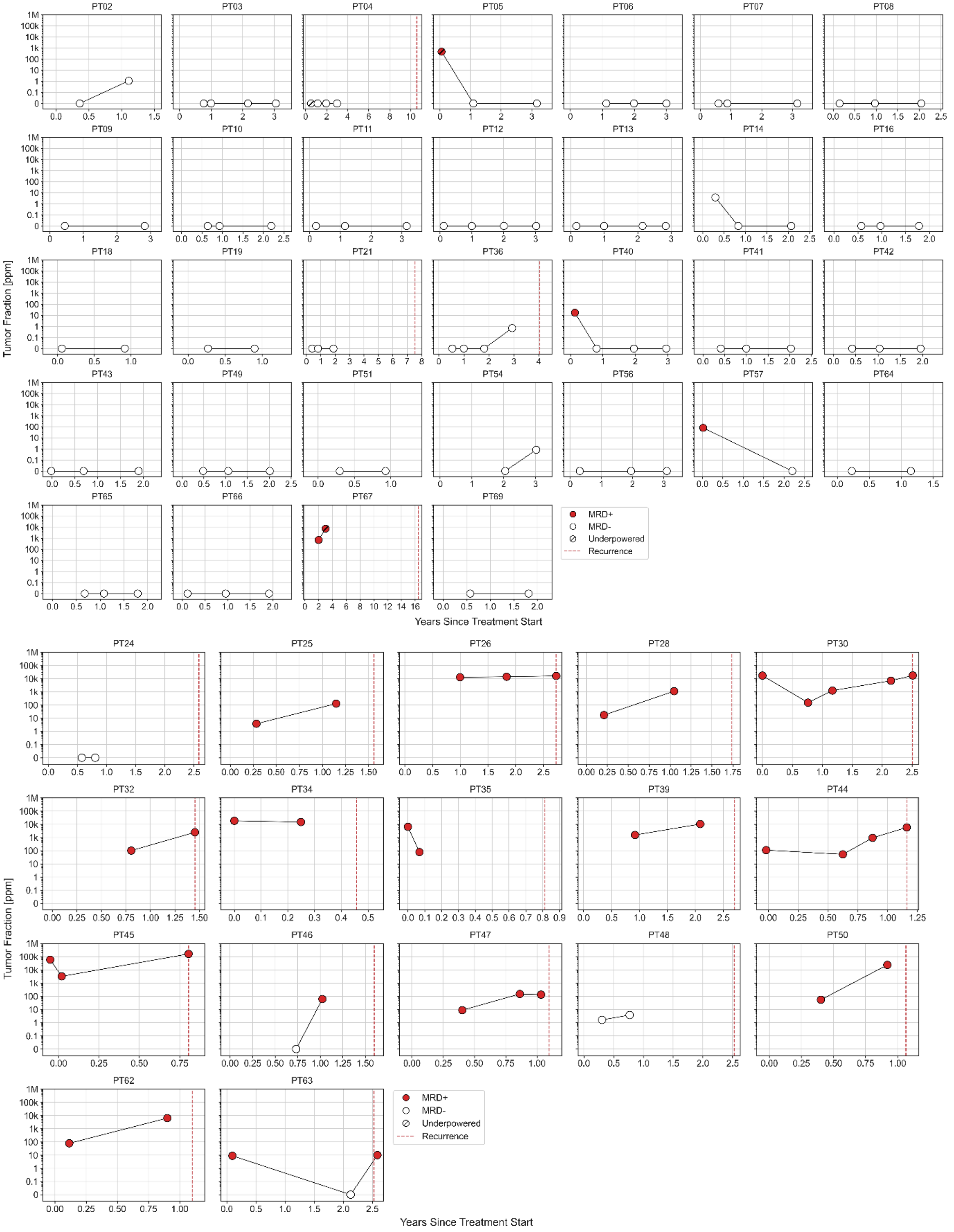

Abbreviations: MRD, minimal residual disease

**Table S1:** MAESTRO-Pool analysis of 149 samples for MRD

| sample_id | patient_id | Tumor_Fraction_ppm | MRD_Detected | LOD95_ppm | ng_input | underpowered | trt_start_to_sample_time_yrs | rwpfs_yrs | rwpfs_event | timepoint |
|---|---|---|---|---|---|---|---|---|---|---|
| PT02_1 | PT02 | 0.00 | FALSE | 6.45 | 15.25 | FALSE | 0.36 | 8.73 | 0 | First-on-tx |
| PT02_2 | PT02 | 0.00 | FALSE | 3.40 | 23.75 | FALSE | 1.11 | 8.73 | 0 | Y1 |
| PT03_1 | PT03 | 0.00 | FALSE | 4.51 | 19.75 | FALSE | 0.77 | 8.68 | 0 | First-on-tx |
| PT03_2 | PT03 | 0.00 | FALSE | 5.71 | 18.00 | FALSE | 1.00 | 8.68 | 0 | Y1 |
| PT03_3 | PT03 | 0.00 | FALSE | 3.48 | 26.25 | FALSE | 2.17 | 8.68 | 0 | Y2 |
| PT03_4 | PT03 | 0.00 | FALSE | 3.29 | 26.25 | FALSE | 3.05 | 8.68 | 0 | Y3 |
| PT04_1 | PT04 | 0.00 | FALSE | 487.67 | 10.34 | TRUE | 0.52 | 10.57 | 1 | First-on-tx |
| PT04_2 | PT04 | 0.00 | FALSE | 6.94 | 18.41 | FALSE | 1.15 | 10.57 | 1 | Y1 |
| PT04_3 | PT04 | 0.00 | FALSE | 58.15 | 6.21 | FALSE | 1.95 | 10.57 | 1 | Y2 |
| PT04_4 | PT04 | 0.00 | FALSE | 12.34 | 11.25 | FALSE | 2.99 | 10.57 | 1 | Y3 |
| PT05_1 | PT05 | 461.31 | TRUE | 438.17 | 13.66 | TRUE | 0.06 | 5.89 | 0 | First-on-tx |
| PT05_2 | PT05 | 0.00 | FALSE | 71.02 | 8.28 | FALSE | 1.10 | 5.89 | 0 | Y1 |
| PT05_3 | PT05 | 0.00 | FALSE | 27.35 | 15.00 | FALSE | 3.17 | 5.89 | 0 | Y3 |
| PT06_1 | PT06 | 0.00 | FALSE | 9.41 | 13.25 | FALSE | 1.13 | 9.05 | 0 | Y1 |
| PT06_2 | PT06 | 0.00 | FALSE | 63.55 | 6.50 | FALSE | 1.99 | 9.05 | 0 | Y2 |
| PT06_3 | PT06 | 0.00 | FALSE | 17.81 | 11.50 | FALSE | 3.01 | 9.05 | 0 | Y3 |
| PT07_1 | PT07 | 0.00 | FALSE | 5.43 | 42.00 | FALSE | 0.59 | 8.55 | 0 | First-on-tx |
| PT07_2 | PT07 | 0.00 | FALSE | 6.74 | 35.49 | FALSE | 0.88 | 8.55 | 0 | Y1 |
| PT07_4 | PT07 | 0.00 | FALSE | 9.45 | 22.00 | FALSE | 3.16 | 8.55 | 0 | Y3 |
| PT08_1 | PT08 | 0.00 | FALSE | 5.32 | 14.25 | FALSE | 0.15 | 8.06 | 0 | First-on-tx |
| PT08_2 | PT08 | 0.00 | FALSE | 6.41 | 14.00 | FALSE | 0.97 | 8.06 | 0 | Y1 |
| PT08_3 | PT08 | 0.00 | FALSE | 5.28 | 12.25 | FALSE | 2.04 | 8.06 | 0 | Y2 |
| PT09_1 | PT09 | 0.00 | FALSE | 7.90 | 28.01 | FALSE | 0.45 | 8.15 | 0 | First-on-tx |
| PT09_3 | PT09 | 0.00 | FALSE | 16.30 | 16.50 | FALSE | 2.83 | 8.15 | 0 | Y3 |
| PT10_1 | PT10 | 0.00 | FALSE | 9.47 | 10.50 | FALSE | 0.64 | 8.59 | 0 | First-on-tx |
| PT10_2 | PT10 | 0.00 | FALSE | 12.50 | 8.25 | FALSE | 0.93 | 8.59 | 0 | Y1 |
| PT10_3 | PT10 | 0.00 | FALSE | 4.80 | 16.00 | FALSE | 2.19 | 8.59 | 0 | Y2 |
| PT11_1 | PT11 | 0.00 | FALSE | 11.55 | 15.25 | FALSE | 0.21 | 7.83 | 0 | First-on-tx |
| PT11_2 | PT11 | 0.00 | FALSE | 11.19 | 15.00 | FALSE | 1.15 | 7.83 | 0 | Y1 |
| PT11_3 | PT11 | 0.00 | FALSE | 10.28 | 15.25 | FALSE | 3.16 | 7.83 | 0 | Y3 |
| PT12_1 | PT12 | 0.00 | FALSE | 8.37 | 19.75 | FALSE | 0.11 | 7.35 | 0 | First-on-tx |
| PT12_2 | PT12 | 0.00 | FALSE | 11.37 | 19.75 | FALSE | 0.99 | 7.35 | 0 | Y1 |
| PT12_3 | PT12 | 0.00 | FALSE | 4.23 | 38.50 | FALSE | 2.00 | 7.35 | 0 | Y2 |
| PT12_4 | PT12 | 0.00 | FALSE | 10.03 | 19.50 | FALSE | 3.02 | 7.35 | 0 | Y3 |

| | | | | | | | | | | |
|---|---|---|---|---|---|---|---|---|---|---|
| PT13_1 | PT13 | 0.00 | FALSE | 4.43 | 27.52 | FALSE | 0.17 | 9.34 | 0 | First-on-tx |
| PT13_2 | PT13 | 0.00 | FALSE | 4.90 | 21.93 | FALSE | 1.00 | 9.34 | 0 | Y1 |
| PT13_3 | PT13 | 0.00 | FALSE | 8.25 | 18.25 | FALSE | 2.15 | 9.34 | 0 | Y2 |
| PT13_4 | PT13 | 0.00 | FALSE | 6.26 | 14.75 | FALSE | 2.84 | 9.34 | 0 | Y3 |
| PT14_1 | PT14 | 0.00 | FALSE | 5.38 | 20.00 | FALSE | 0.29 | 5.83 | 0 | First-on-tx |
| PT14_2 | PT14 | 0.00 | FALSE | 9.44 | 12.00 | FALSE | 0.83 | 5.83 | 0 | Y1 |
| PT14_3 | PT14 | 0.00 | FALSE | 5.46 | 20.00 | FALSE | 2.07 | 5.83 | 0 | Y2 |
| PT16_1 | PT16 | 0.00 | FALSE | 11.37 | 29.25 | FALSE | 0.57 | 5.89 | 0 | First-on-tx |
| PT16_2 | PT16 | 0.00 | FALSE | 22.21 | 17.00 | FALSE | 0.97 | 5.89 | 0 | Y1 |
| PT16_3 | PT16 | 0.00 | FALSE | 6.17 | 40.50 | FALSE | 1.78 | 5.89 | 0 | Y2 |
| PT17_1 | PT17 | 0.00 | FALSE | 49.10 | 15.25 | FALSE | 0.82 | 6.16 | 0 | Y1 |
| PT18_1 | PT18 | 0.00 | FALSE | 35.22 | 23.75 | FALSE | 0.06 | 5.60 | 0 | First-on-tx |
| PT18_2 | PT18 | 0.00 | FALSE | 27.36 | 27.00 | FALSE | 0.92 | 5.60 | 0 | Y1 |
| PT19_1 | PT19 | 0.00 | FALSE | 2.37 | 62.74 | FALSE | 0.27 | 5.65 | 0 | First-on-tx |
| PT19_2 | PT19 | 0.00 | FALSE | 13.17 | 10.75 | FALSE | 0.90 | 5.65 | 0 | Y1 |
| PT21_1 | PT21 | 0.00 | FALSE | 22.20 | 13.75 | FALSE | 0.40 | 7.52 | 1 | First-on-tx |
| PT21_2 | PT21 | 0.00 | FALSE | 9.99 | 23.75 | FALSE | 0.81 | 7.52 | 1 | Y1 |
| PT21_3 | PT21 | 0.00 | FALSE | 12.24 | 19.75 | FALSE | 1.88 | 7.52 | 1 | Y2 |
| PT24_1 | PT24 | 0.00 | FALSE | 13.59 | 9.52 | FALSE | 0.57 | 2.58 | 1 | First-on-tx |
| PT24_2 | PT24 | 0.00 | FALSE | 98.22 | 18.00 | FALSE | 0.80 | 2.58 | 1 | Y1 |
| PT25_1 | PT25 | 3.79 | TRUE | 5.27 | 26.69 | FALSE | 0.28 | 1.56 | 1 | First-on-tx |
| PT25_2 | PT25 | 127.22 | TRUE | 31.77 | 8.25 | FALSE | 1.15 | 1.56 | 1 | Y1 |
| PT26_1 | PT26 | 12859.57 | TRUE | 2.72 | 41.50 | FALSE | 0.99 | 2.73 | 1 | Y1 |
| PT26_2 | PT26 | 13987.75 | TRUE | 6.66 | 17.25 | FALSE | 1.83 | 2.73 | 1 | Y2 |
| PT26_3 | PT26 | 16040.09 | TRUE | 2.10 | 48.75 | FALSE | 2.73 | 2.73 | 1 | PD |
| PT27_2 | PT27 | 0.00 | FALSE | 7.80 | 23.25 | FALSE | 1.10 | 2.95 | 1 | Y1 |
| PT28_1 | PT28 | 17.23 | TRUE | 2.52 | 34.75 | FALSE | 0.21 | 1.74 | 1 | First-on-tx |
| PT28_2 | PT28 | 1122.81 | TRUE | 3.02 | 33.25 | FALSE | 1.05 | 1.74 | 1 | Y1 |
| PT29_1 | PT29 | 25.69 | TRUE | 1.28 | 100.00 | FALSE | 0.99 | 2.02 | 1 | Y1 |
| PT30_1 | PT30 | 17025.95 | TRUE | 5.43 | 15.50 | FALSE | 0.00 | 2.51 | 1 | Baseline |
| PT30_2 | PT30 | 150.81 | TRUE | 5.34 | 14.50 | FALSE | 0.76 | 2.51 | 1 | First-on-tx |
| PT30_3 | PT30 | 1247.85 | TRUE | 5.16 | 12.00 | FALSE | 1.17 | 2.51 | 1 | Y1 |
| PT30_4 | PT30 | 6967.90 | TRUE | 6.71 | 14.00 | FALSE | 2.15 | 2.51 | 1 | Y2 |
| PT30_5 | PT30 | 17527.25 | TRUE | 8.25 | 11.50 | FALSE | 2.51 | 2.51 | 1 | PD |

| | | | | | | | | | | |
|---|---|---|---|---|---|---|---|---|---|---|
| PT31_1 | PT31 | 6113.68 | TRUE | 2.66 | 25.25 | FALSE | 0.92 | 1.10 | 1 | Y1 |
| PT32_1 | PT32 | 101.14 | TRUE | 10.91 | 24.50 | FALSE | 0.80 | 1.45 | 1 | Y1 |
| PT32_2 | PT32 | 2467.63 | TRUE | 9.57 | 13.50 | FALSE | 1.45 | 1.45 | 1 | PD |
| PT33_1 | PT33 | 1652.55 | TRUE | 3.50 | 46.00 | FALSE | 0.00 | 0.48 | 1 | Baseline |
| PT34_1 | PT34 | 18438.37 | TRUE | 1.09 | 100.00 | FALSE | 0.00 | 0.46 | 1 | Baseline |
| PT34_2 | PT34 | 14858.07 | TRUE | 2.69 | 22.50 | FALSE | 0.25 | 0.46 | 1 | First-on-tx |
| PT35_1 | PT35 | 6475.65 | TRUE | 5.17 | 27.50 | FALSE | 0.00 | 0.81 | 1 | Baseline |
| PT35_2 | PT35 | 77.80 | TRUE | 6.12 | 26.89 | FALSE | 0.07 | 0.81 | 1 | First-on-tx |
| PT36_1 | PT36 | 0.00 | FALSE | 6.10 | 12.41 | FALSE | 0.54 | 4.04 | 1 | First-on-tx |
| PT36_2 | PT36 | 0.00 | FALSE | 2.88 | 36.09 | FALSE | 1.00 | 4.04 | 1 | Y1 |
| PT36_3 | PT36 | 0.00 | FALSE | 3.62 | 20.75 | FALSE | 1.82 | 4.04 | 1 | Y2 |
| PT36_4 | PT36 | 0.00 | FALSE | 1.58 | 71.74 | FALSE | 2.93 | 4.04 | 1 | Y3 |
| PT37_1 | PT37 | 127.74 | TRUE | 303.85 | 10.76 | TRUE | 0.99 | 2.69 | 1 | Y1 |
| PT38_1 | PT38 | 0.00 | FALSE | 13.55 | 17.00 | FALSE | 0.96 | 4.32 | 1 | Y1 |
| PT39_1 | PT39 | 1547.35 | TRUE | 20.52 | 27.50 | FALSE | 0.92 | 2.70 | 1 | Y1 |
| PT39_2 | PT39 | 10415.40 | TRUE | 72.18 | 15.00 | FALSE | 2.09 | 2.70 | 1 | Y2 |
| PT40_1 | PT40 | 16.90 | TRUE | 20.05 | 17.00 | FALSE | 0.14 | 5.50 | 0 | First-on-tx |
| PT40_2 | PT40 | 0.00 | FALSE | 47.81 | 9.00 | FALSE | 0.81 | 5.50 | 0 | Y1 |
| PT40_3 | PT40 | 0.00 | FALSE | 18.07 | 16.50 | FALSE | 1.96 | 5.50 | 0 | Y2 |
| PT40_4 | PT40 | 0.00 | FALSE | 19.43 | 18.75 | FALSE | 2.95 | 5.50 | 0 | Y3 |
| PT41_1 | PT41 | 0.00 | FALSE | 2.49 | 37.50 | FALSE | 0.41 | 4.81 | 0 | First-on-tx |
| PT41_2 | PT41 | 0.00 | FALSE | 6.04 | 21.00 | FALSE | 1.00 | 4.81 | 0 | Y1 |
| PT41_3 | PT41 | 0.00 | FALSE | 2.71 | 31.50 | FALSE | 2.03 | 4.81 | 0 | Y2 |
| PT42_1 | PT42 | 0.00 | FALSE | 17.86 | 15.00 | FALSE | 0.42 | 4.54 | 0 | First-on-tx |
| PT42_2 | PT42 | 0.00 | FALSE | 5.65 | 27.50 | FALSE | 1.03 | 4.54 | 0 | Y1 |
| PT42_3 | PT42 | 0.00 | FALSE | 5.40 | 26.50 | FALSE | 1.95 | 4.54 | 0 | Y2 |
| PT43_1 | PT43 | 0.00 | FALSE | 3.44 | 16.75 | FALSE | -0.02 | 8.62 | 0 | Baseline |
| PT43_2 | PT43 | 0.00 | FALSE | 17.56 | 7.75 | FALSE | 0.69 | 8.62 | 0 | First-on-tx |
| PT43_4 | PT43 | 0.00 | FALSE | 5.48 | 13.50 | FALSE | 1.90 | 8.62 | 0 | Y2 |
| PT44_1 | PT44 | 109.68 | TRUE | 8.83 | 32.25 | FALSE | -0.02 | 1.16 | 1 | Baseline |
| PT44_2 | PT44 | 52.56 | TRUE | 13.13 | 18.25 | FALSE | 0.62 | 1.16 | 1 | First-on-tx |
| PT44_3 | PT44 | 936.47 | TRUE | 37.05 | 16.25 | FALSE | 0.87 | 1.16 | 1 | Y1 |
| PT44_4 | PT44 | 5798.51 | TRUE | 16.74 | 25.25 | FALSE | 1.16 | 1.16 | 1 | PD |

| | | | | | | | | | | |
|---|---|---|---|---|---|---|---|---|---|---|
| PT45_1 | PT45 | 59814.48 | TRUE | 6.72 | 25.50 | FALSE | -0.05 | 0.80 | 1 | Baseline |
| PT45_2 | PT45 | 3266.53 | TRUE | 5.43 | 42.00 | FALSE | 0.02 | 0.80 | 1 | First-on-tx |
| PT45_3 | PT45 | 164068.22 | TRUE | 3.74 | 46.25 | FALSE | 0.80 | 0.80 | 1 | PD |
| PT46_1 | PT46 | 0.00 | FALSE | 42.81 | 12.75 | FALSE | 0.73 | 1.60 | 1 | First-on-tx |
| PT46_2 | PT46 | 61.56 | TRUE | 14.61 | 21.25 | FALSE | 1.02 | 1.60 | 1 | Y1 |
| PT47_1 | PT47 | 8.86 | TRUE | 13.57 | 19.00 | FALSE | 0.40 | 1.10 | 1 | First-on-tx |
| PT47_2 | PT47 | 149.95 | TRUE | 11.12 | 20.00 | FALSE | 0.86 | 1.10 | 1 | Y1 |
| PT47_3 | PT47 | 132.75 | TRUE | 18.54 | 14.00 | FALSE | 1.03 | 1.10 | 1 | PD |
| PT48_1 | PT48 | 0.00 | FALSE | 4.95 | 22.75 | FALSE | 0.30 | 2.53 | 1 | First-on-tx |
| PT48_2 | PT48 | 0.00 | FALSE | 5.51 | 17.75 | FALSE | 0.77 | 2.53 | 1 | Y1 |
| PT49_1 | PT49 | 0.00 | FALSE | 5.92 | 13.00 | FALSE | 0.48 | 8.42 | 0 | First-on-tx |
| PT49_2 | PT49 | 0.00 | FALSE | 4.96 | 41.80 | FALSE | 1.06 | 8.42 | 0 | Y1 |
| PT49_3 | PT49 | 0.00 | FALSE | 5.40 | 17.25 | FALSE | 2.01 | 8.42 | 0 | Y2 |
| PT50_1 | PT50 | 54.63 | TRUE | 11.79 | 22.00 | FALSE | 0.40 | 1.07 | 1 | First-on-tx |
| PT50_2 | PT50 | 23963.50 | TRUE | 9.66 | 26.50 | FALSE | 0.92 | 1.07 | 1 | Y1 |
| PT51_1 | PT51 | 0.00 | FALSE | 5.68 | 22.50 | FALSE | 0.30 | 5.59 | 0 | First-on-tx |
| PT51_2 | PT51 | 0.00 | FALSE | 19.33 | 9.25 | FALSE | 0.93 | 5.59 | 0 | Y1 |
| PT53_1 | PT53 | 13.99 | TRUE | 4.00 | 19.45 | FALSE | 2.97 | 5.74 | 1 | Y3 |
| PT54_2 | PT54 | 0.00 | FALSE | 3.12 | 67.74 | FALSE | 2.05 | 5.33 | 0 | Y2 |
| PT54_3 | PT54 | 0.00 | FALSE | 3.12 | 100.00 | FALSE | 3.03 | 5.33 | 0 | Y3 |
| PT55_1 | PT55 | 11.90 | TRUE | 4.41 | 32.50 | FALSE | 3.03 | 6.59 | 1 | Y3 |
| PT56_1 | PT56 | 0.00 | FALSE | 4.18 | 41.50 | FALSE | 0.31 | 6.09 | 0 | First-on-tx |
| PT56_2 | PT56 | 0.00 | FALSE | 6.01 | 31.00 | FALSE | 1.95 | 6.09 | 0 | Y2 |
| PT56_3 | PT56 | 0.00 | FALSE | 4.64 | 33.50 | FALSE | 3.10 | 6.09 | 0 | Y3 |
| PT57_1 | PT57 | 78.57 | TRUE | 6.51 | 15.00 | FALSE | 0.02 | 6.44 | 0 | First-on-tx |
| PT57_2 | PT57 | 0.00 | FALSE | 6.69 | 16.75 | FALSE | 2.21 | 6.44 | 0 | Y2 |
| PT58_1 | PT58 | 0.00 | FALSE | 5.03 | 23.75 | FALSE | 1.91 | 5.30 | 0 | Y2 |
| PT59_1 | PT59 | 0.00 | FALSE | 4.79 | 22.50 | FALSE | 2.02 | 4.97 | 0 | Y2 |
| PT61_1 | PT61 | 3155.91 | TRUE | 3.53 | 45.72 | FALSE | 1.97 | 2.15 | 1 | Y2 |
| PT62_1 | PT62 | 78.69 | TRUE | 1.20 | 44.75 | FALSE | 0.11 | 1.10 | 1 | First-on-tx |
| PT62_2 | PT62 | 6416.75 | TRUE | 1.56 | 40.00 | FALSE | 0.90 | 1.10 | 1 | Y1 |
| PT63_1 | PT63 | 8.86 | TRUE | 0.77 | 100.00 | FALSE | 0.10 | 2.53 | 1 | First-on-tx |
| PT63_2 | PT63 | 0.00 | FALSE | 1.41 | 39.25 | FALSE | 2.13 | 2.53 | 1 | Y2 |

| | | | | | | | | | | |
|---|---|---|---|---|---|---|---|---|---|---|
| PT63_3 | PT63 | 10.00 | TRUE | 1.79 | 30.00 | FALSE | 2.59 | 2.53 | 1 | PD |
| PT64_1 | PT64 | 0.00 | FALSE | 5.91 | 29.75 | FALSE | 0.23 | 4.52 | 0 | First-on-tx |
| PT64_2 | PT64 | 0.00 | FALSE | 14.97 | 12.75 | FALSE | 1.15 | 4.52 | 0 | Y1 |
| PT65_1 | PT65 | 0.00 | FALSE | 5.97 | 34.50 | FALSE | 0.68 | 4.36 | 0 | First-on-tx |
| PT65_2 | PT65 | 0.00 | FALSE | 16.98 | 15.25 | FALSE | 1.08 | 4.36 | 0 | Y1 |
| PT65_3 | PT65 | 0.00 | FALSE | 2.92 | 29.50 | FALSE | 1.79 | 4.36 | 0 | Y2 |
| PT66_1 | PT66 | 0.00 | FALSE | 3.41 | 35.00 | FALSE | 0.11 | 4.03 | 0 | First-on-tx |
| PT66_2 | PT66 | 0.00 | FALSE | 3.64 | 35.00 | FALSE | 0.96 | 4.03 | 0 | Y1 |
| PT66_3 | PT66 | 0.00 | FALSE | 3.96 | 34.00 | FALSE | 1.92 | 4.03 | 0 | Y2 |
| PT67_1 | PT67 | 677.03 | TRUE | 54.46 | 22.55 | FALSE | 1.99 | 16.48 | 1 | Y2 |
| PT67_2 | PT67 | 7152.51 | TRUE | 129.55 | 14.48 | TRUE | 3.01 | 16.48 | 1 | Y3 |
| PT68_1 | PT68 | 0.00 | FALSE | 10.38 | 30.50 | FALSE | 2.95 | 10.29 | 0 | Y3 |
| PT69_1 | PT69 | 0.00 | FALSE | 7.66 | 27.25 | FALSE | 0.57 | 7.47 | 0 | First-on-tx |
| PT69_2 | PT69 | 0.00 | FALSE | 55.17 | 9.75 | FALSE | 1.82 | 7.47 | 0 | Y2 |
| PT70_1 | PT70 | 0.00 | FALSE | 3.15 | 27.00 | FALSE | 3.17 | 5.81 | 0 | Y3 |

| Column Name | Description |
| --- | --- |
| sample_id | ID associated with sample under the same patient |
| patient_id | Patient ID |
| Tumor_Fraction_ppm | Tumor fraction |
| MRD_Detected | Minimal Residual Disease status |
| LOD95_ppm | Limit of detection for sample |
| ng_input | Mass input into library construction (ng) |
| underpowered | The LOD95 is **greater** than 100ppm |
| trt_start_to_sample_time_yrs | Time interval from 1L treatment start to blood sampling (years) |
| rwpfs_yrs | Time interval from 1L treatment start to rwPFS event or last follow up (years) |
| rwpfs_event | rwPFS event: yes = 1; no =0 |
| timepoint | Key timepoint category (baseline, first on therapy [first-on-tx], year 1 [Y1], year 2 [Y2], year 3 [Y3], at progression [PD]) |

**Table S2.** Clinicopathological features according to MRD status at Y1.

| | MRD-positive (N=9) | MRD-negative (N=30) | Total (N=39) | p value |
|---|---|---|---|---|
| **Stage at Initial Diagnosis** | | | | 0.238 |
| I to III | 5 (55.6%) | 9 (30.0%) | 14 (35.9%) | |
| IV | 4 (44.4%) | 21 (70.0%) | 25 (64.1%) | |
| **HER2 Status** | | | | 0.355 |
| 0-2+/FISH positive | 3 (33.3%) | 5 (16.7%) | 8 (20.5%) | |
| 3+ | 6 (66.7%) | 25 (83.3%) | 31 (79.5%) | |
| **Estrogen Receptor Status** | | | | 0.704 |
| Positive | 3 (33.3%) | 14 (46.7%) | 17 (43.6%) | |
| Negative | 6 (66.7%) | 16 (53.3%) | 22 (56.4%) | |
| **Number of Disease Sites at MBC diagnosis** | | | | 1.000 |
| 1-2 | 8 (88.9%) | 24 (80.0%) | 32 (82.1%) | |
| $\geqslant 3$ | 1 (11.1%) | 6 (20.0%) | 7 (17.9%) | |
| **Visceral Disease at MBC diagnosis** | | | | 0.406 |
| Yes | 6 (66.7%) | 24 (80.0%) | 30 (76.9%) | |
| No | 3 (33.3%) | 6 (20.0%) | 9 (23.1%) | |

Abbreviations: MRD, minimal residual disease; Y, year; FISH, Fluorescence In Situ Hybridization; MBC, metastatic breast cancer
